# Exposure to landscape fire smoke extremely reduced birthweight in low- and middle-income countries

**DOI:** 10.1101/2021.04.27.21256225

**Authors:** Jiajianghui Li, Tianjia Guan, Qian Guo, Guannan Geng, Huiyu Wang, Fuyu Guo, Jiwei Li, Tao Xue

## Abstract

Landscape fire smoke (LFS) has been associated with reduced birthweight, but evidence from low- and middle-income countries (LMICs) is rare. Here, we present a sibling-matched case– control study of 227,948 newborns to identify an association between fire-sourced fine particulate matter (PM_2.5_) and birthweight in 54 LMICs from 2000 to 2014.

We selected mothers from the geocoded Demographic and Health Survey with at least two children and valid birthweight records. Newborns affiliated with the same mother were defined as a family group. Gestational exposure to LFS was assessed in each newborn using the concentration of fire-sourced PM_2.5_. We determined the associations of the within-group variations in LFS exposure with birthweight differences between matched siblings using a fixed-effects regression model. Additionally, we analyzed the binary outcomes of low birthweight (LBW) or very low birthweight (VLBW).

According to fully adjusted models, a 1 µg/m^3^ increase in the concentration of fire-sourced PM_2.5_ was significantly associated with a 2.17 g (95% confidence interval [CI]: 0.56–3.77) reduction in birthweight, a 2.80% (95% CI: 0.97–4.66) increase in LBW risk, and an 11.68% (95% CI: 3.59–20.40) increase in VLBW risk.

Our findings indicate that gestational exposure to LFS harms maternal health.

## Introduction

The natural cycle of landscape fires (e.g., wildfires, tropical deforestation fires, and agricultural biomass burning) plays an important role in maintaining the terrestrial ecosystem. Yet, landscape fire smoke (LFS) triggers a costly and growing global public health problem^1^. The frequency and intensity of landscape fire events have tended to increase^2,3^, driven by interactions between human activities (e.g., slash and slave agriculture) and climate change^4^. Most emissions originate from fires located in tropical rainforests and savannas, where they cause recurrent episodes of severe ambient pollution that affect mainly low- and middle-income countries (LMICs)^1^. Worldwide concern regarding large-scale natural forest burning has sparked increasing interest in the harmful effects of LFS exposure on human health.

LFS is composed of hundreds of combustion products, such as carbon monoxide, nitrogen oxides, and polycyclic aromatic hydrocarbons^5^. Particulate matter with an aerodynamic diameter ≤ 2.5 μm (PM_2.5_) is one of the most widely studied pollutants derived from LFS. The 24-h average PM_2.5_ concentrations on burning days can be many times higher than those on normal days, and the peak level of ambient exposure can persist for several weeks^6^. Several epidemiological studies have shown adverse effects of LFS exposure on human health. The majority of the study outcomes involved morbidity and mortality caused by acute respiratory and cardiovascular diseases, particularly among vulnerable individuals, such as children and the elderly^7-9^. However, the health impacts of LFS on susceptible pregnant women, another highly vulnerable group due to gestation-related physiological changes, such as an increased breathing rate during pregnancy^10^, are rarely discussed. Previous studies have shown that gestational exposure to air pollutants, including those derived from LPS and other emissions, are associated with abnormal placental vascular function and, consequently, low birthweight (LBW)^11^, preterm birth^12^, and small for gestational age^13^. Exposure to air pollution induces subclinical disorders, such as inflammation, oxidative stress, increased blood viscosity, mitochondrial methylation, and hypoxia^14,15^, which renders the association between LFS and adverse maternal outcomes biologically plausible.

Birth weight comprehensively reflects the intrauterine environment quality, fetal growth, and maternal nutritional status. Therefore, LBW is a global public health problem. LBW neonates are at higher risk for a range of diseases in later life (e.g., early growth retardation and adulthood cardiorespiratory diseases) compared with normal newborns^16,17^. The estimated worldwide prevalence of LBW was 14.6% (95% confidence interval [CI] 12.4–17.1) in 2015, compared with 17.5% (95% CI 14.1–21.3) in 2000^18^. Although some progress has been made in reducing LBW globally, the average annual reduction rate is insufficient to achieve the 2025 WHO global target^19^. Additionally, the relevant burden of LBW is unequally distributed in the world, as 91% of LBW infants are from LMICs, particularly from southern Asia and sub-Saharan Africa^18^.

All risk factors for adverse birth defects should be screened thoroughly to protect maternal and infant health. In addition to conventional factors, such as nutrition and maternal condition, environmental factors, including ambient PM_2.5_ exposure, also contribute to a considerable global burden of adverse birth outcomes^20^. Although a growing number of studies have suggested the deleterious effects of urban air pollution on LBW^21,22^, little is known about fire-sourced PM_2.5_. Furthermore, studies concerning the association between LFS and birthweight have focused primarily on high-income countries, and their conclusions have been inconsistent^23-26^. As most landscape fires occur in LMICs (Figure 1), studies to assess the relevant health impacts of LFS among residents of such countries are warranted. Hence, we explored the association between birthweight and fire-sourced PM_2.5_ in 54 LMICs using a sibling-matched case–control study from 2000 to 2014, which was based on a series of global surveys.

**Figure 1.**
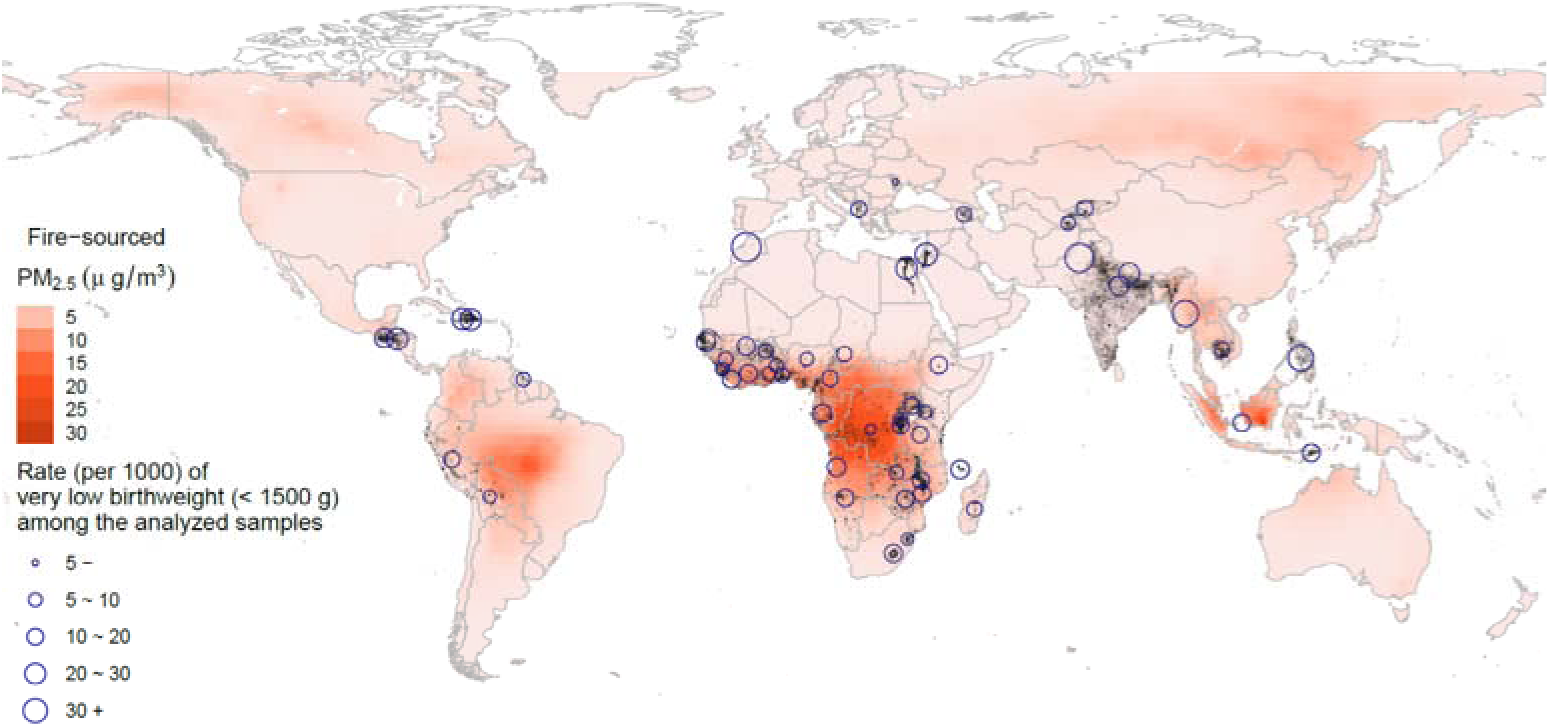
Distribution of the fire-sourced PM_2.5_ concentrations (average for 2000–2014) and the locations of the analyzed samples (grey dots). The circles represent the rate of very-low-birthweight births among the analyzed samples by country.

## Methods

### Study population

The birthweight data and relevant individual variables were extracted from 113 individual Demographic and Health Surveys (DHSs) representing 54 LMICs from 2000 to 2014 (Figure 1). The DHSs were conducted by the US Agency for International Development to characterize the demography and health of more than 90 developing countries. DHSs are usually conducted every 5 years in each country, using a two-stage sampling design. Information on demographic factors, socioeconomic factors, and health status was collected from a nationally representative probability sample of households. Well-trained fieldworkers conducted the surveys using uniformly designed questionnaires in the native language. The primary sampling units (e.g., villages or blocks) have begun to be geocoded in recent waves using global positioning systems (GPSs). The GPS-enhanced DHSs were matched with environmental exposure and thus included in our analyses. Please refer to the open-access website for detailed information on the DHS database: https://www.dhsprogram.com/.

All females aged 15–49 years within each selected household were eligible respondents of the survey. The health outcome analyzed in this study was birthweight. The DHSs asked the screened mothers to recall the birthweight of all children born during the 5 years leading up to the survey date. To characterize different levels of reduced birthweight, we examined two binary outcomes: LBW (birthweight < 2,500 g) and very LBW (VLBW, birthweight < 1,500 g). Other covariates, such as maternal age and birth time (in months), were extracted from the relevant questionnaires.

### Exposure assessment

We assessed ambient exposure to LFS as the PM_2.5_ concentration attributable to landscape fires (hereafter, defined as fire-sourced PM_2.5_), referring to a previous study^27^. The fire-sourced PM_2.5_ concentrations were estimated using the GEOS-Chem chemical transport model (CTM). The detailed settings of the GEOS-Chem are provided in the supplemental text.

The fire-sourced PM_2.5_ was derived from a comparison between two GEOS-Chem model runs. The two models were identical except for switching on and off fire emissions. We quantified the fraction (*ρ*_*m*_) of fire-attributed PM_2.5_ using the following equation:

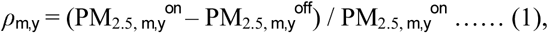

where the superscripts *on* and *off* denote the two simulations switching on and off fire emissions, respectively; and the subscripts *m* and *y* are month and year indicators, respectively.

Additionally, considering the potential errors in the CTM procedure^28^, the bias correction approach has been widely applied to improve CTM performance in exposure assessments using ground-surface observational data^29^. We utilized a well-developed and widely utilized product of satellite-based PM_2.5_ estimates^30^ (PM_2.5, *y*_ ^*satellite*^) in the bias correction. This product has a spatial resolution of 0.05° × 0.05° and was publicly available from 1998 to 2018 as the gridded annual mean (http://fizz.phys.dal.ca/~atmos/martin/?page_id=140#V4.GL.03). We prepared the GEOS-Chem simulations (ρ_*m,y*_ and PM_2.5, *m,y*_^*on*^) in the same 0.05° × 0.05° grid using an inversed distance-weighted downscaling approach before bias correction. The bias correction rate (η_*y*_) was calculated as

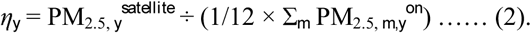

The monthly exposure indicator of fire-sourced or non-fire-sourced PM_2.5_ was finally derived as

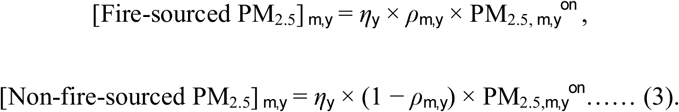

We utilized the non-fire-sourced PM_2.5_ as a potential confounder in the subsequent epidemiological analysis. We also collected other environmental variables, including ambient temperature and humidity data, from an assimilated dataset called Modern-Era Retrospective analysis for Research and Applications, version 2 (please refer to the supplemental text for more details). The variables were downloaded as monthly gridded values and prepared in the 0.05° × 0.05° grid in the same way as we processed the GEOS-Chem outputs.

To understand the associations between birthweight and environmental variables, we focused on exposure during gestation. As the DHSs did not record the specific duration of gestation, we utilized the 9-month average preceding the survey month as exposure time during pregnancy.

### Study design

To evaluate the association between fire-sourced PM_2.5_ and birthweight, we applied a sibling-matched case–control study, similar to our previous work^31^. We defined the descendants of the same mother as a family group of matched siblings and compared birthweights within each group. The family-level baseline birthweight (defined as the mean birthweight within each group) can be affected by many complex factors, including genetics, socioeconomic position, and quality of local medical services, which can be difficult to measure. We matched the mothers to control for those unmeasured confounders in the study design. A sibling-matched case–control design is a simple, robust, and cost-efficient method, particularly for large-population data with potential heterogeneity^32,33^.

We extracted the females with at as least two valid records of newborns from all available individual DHSs. Each valid record was defined using the following inclusion criteria: (1) a valid birthweight, (2) a record of the GPS coordinates, (3) a valid birthdate, and (4) valid environmental exposure values (some of the satellite-based PM_2.5_ estimates were not available for some locations, such as small islands). Finally, our dataset assembled 113 individual surveys from 54 countries (Figure 1) and included 227,948 newborns classified into 109,137 groups according to their mothers.

### Statistical analyses

According to the sibling-matched method, we used a fixed-effects regression model to evaluate the associations between fire-sourced PM_2.5_ and LBW or VLBW. The model was specified as

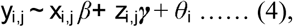

where the subscripts *i* and *j* denote the indices of the mother-affiliated group and child, respectively; *x*_*i,j*_ denotes the concentration of fire-sourced PM_2.5_; ***z***_*i,j*_ denotes the adjusted covariates; *y*_*i,j*_ denotes the outcome variable; and *θ*_*i*_ is a nuisance parameter for the fixed-effect to control for group-specific unmeasured confounders. The model was specified as Gaussian regression when setting *y*_*i,j*_ as the birthweight and as logit regression when setting *y*_*i,j*_ as the logit-transformed probability of LBW or VLBW. The adjusted covariates included maternal age; child sex; multiple births; non-fire-sourced PM_2.5_; spline terms of birth order (with 5 degrees of freedom [DF]), of temperature (3 DF), of humidity (3 DF), of the calendar year (5 DF), and of the monthly index (4 DF); and a spatiotemporal effect parametrized by the interaction between country and calendar year. The nonlinear effects of month and year controlled for the seasonal periodic variation and long-term trend in child health, respectively. The spatiotemporal effect captured country-specific trends. The association between birthweight and LFS was evaluated by the regression coefficient *β* for 1 μg/m^3^ increases in fire-sourced PM_2.5_. The association with LBW or VLBW was evaluated by excess risk [= (1 – exp(*β*)) × 100%] for a 1 μg/m^3^ increase in fire-sourced PM_2.5_. All environmental exposures were quantified using the same time window. The major time window was the 9 months before birth. We also calculated the alternative exposure indicator as an average during the 3 or 6 months preceding birth.

In sensitivity analyses, we first derived the lag-distributed model to explore how the association varied during the exposure time window. Next, subpopulation-specified associations were estimated using interaction analyses to detect potential heterogeneity. Third, we developed nonlinear associations by replacing the linear term of fire-sourced PM_2.5_ using a set of thin-plate spline functions. Fourth, to examine potential recall bias, we estimated the associations by strata of the durations from birth to survey time. Fifth, to test whether the estimated associations were attributable to LFS exposure or other direct damage related to landscape fires (e.g., destroying human habitats), we derived an indicator for the transported fire-sourced PM_2.5_, according to a satellite image of the burned area (MCD64A1, https://lpdaacsvc.cr.usgs.gov/appeears/). The fire-sourced PM_2.5_ at the pixels with the zero-burned area during gestation were defined as smoke transported from other locations. The associations with transported fire-sourced PM_2.5_ could be indicative of the effect of LFS exposure via the inhalation pathway.

Family-level baseline birthweight and the correlated gestation length can be determined by factors such as genetics and socioeconomic level, which may also affect the association between the within-group change in birthweight and LFS exposure. The fixed-effects regression models actually infer an association by examining the coherence between derivation of individual-level birthweight from the family-level mean (i.e., the expected birthweight given a specific family) and the corresponding exposure. To show that, we defined the family-level baseline birthweight as *y*_*i*_. Equation (4) was equivalently transformed as Δ*y*_*i,j*_ = (*y*_*i,j*_ − *y_*i*_) *∼ x*_*i,j*_ *β* +* ***z***_*i,j*_*γ* + (*θ*_*i*_ − *y*_*i*_), where (*θ*_*i*_ − *y*_*i*_) is viewed as a new nuisance parameter *θ* ^*^. The nature of fixed-effects model made it possible to explore how the effect of fire-sourced PM_2.5_ varied according to family-level mean birthweight. Accordingly, we derived a baseline-varying association model as follows:

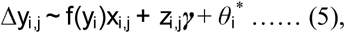

where *f(y*_*i*_) denotes the baseline-varying coefficient and is parametrized by a set of spline functions.

All analyses were performed in R (version 4.0.2) (The R Foundation for Statistical Computing, Vienna, Austria). Inference for the fixed-effects models was performed using the *fixest* package. The lag-distributed effect was parametrized using the *dlnm* package, and the thin-plate splines for the nonlinear association and baseline-varying effect were parametrized using the *mgcv* package.

## Results

### Descriptive summary

Among the 227,948 samples analyzed from 54 LMICs, there were 109,137 groups of siblings, who were matched to their mothers. Each group had an average of 2.13 (standard deviation [SD]: 0.36) samples. Approximately half of the samples were from Sub-Saharan Africa and one-quarter from South Asia. The mean birthweight was 3,082 (SD: 724) g. There were 31,854 LBW births and 2,912 VLBW births. More statistics on the analyzed samples are shown in Supplemental Table S1.

The spatial distribution of the long-term average fire-sourced PM_2.5_ concentrations is displayed in Figure 1. Our modeling results indicate that the hotspots of LFS include regions of the Congo rainforest basin, Amazon rainforest basin, Indonesian forests, Siberia forests, and other forest areas. Our analyzed samples included newborns from the first three regions. Among our samples, the average concentration of full gestational exposure to fire-sourced PM_2.5_ was 4.29 (SD: 5.53) µg/m^3^. A detailed summary of the environmental exposures is presented in Table S1.

Gestational exposure to fire-sourced PM_2.5_ varied widely between family groups; it was higher in those living closer to hotspot regions (e.g., forests). Approximately 92% of the variation in fire-sourced PM_2.5_ exposure was between family groups, and 8% was within family groups. For the total variation in birthweight, 71.7% was between groups and 28.3% within groups (Table 1). The within-or between-group variations reflected temporal or spatial changes in those variables, respectively. Birthweight and fire-sourced PM_2.5_ were negatively correlated in the temporal dimension but positively correlated in the spatial dimension. As the spatial patterns in birthweight and fire-sourced PM_2.5_ were determined by geographic, socioeconomic, and cultural factors, the between-group correlations may have been confounded by their common determiners, such as urbanization level. The complex correlations between birthweight and LFS suggest that statistical inference of their associations should be cautiously determined based on well-designed epidemiological analysis.

**Table 1.** Between- or within-group variations in birthweight and gestational exposure to fire-sourced PM_2.5_ and their correlations.

|  | Standard deviation (% of total variance) |  | Correlation (R) |
| --- | --- | --- | --- |
|  | Birthweight | Fire-sourced PM<sub>2.5</sub> |  |
| Total | 724 (100%) | 5.53 (100%) | 0.1594 |
| Between groups of matched siblings | 613 (71.7%) | 5.30 (91.7%) | 0.1973 |
| Within groups of matched siblings | 386 (28.3%) | 1.59 (8.3%) | -0.0035 |

### Association between LFS and birthweight

Although the fully adjusted model indicated that a 1 µg/m^3^ increase in exposure to fire-sourced PM_2.5_ during the 9 months before birth was significantly associated with a 2.17 g (95% CI: 0.56–3.77) birthweight reduction, the association was sensitive to differences in the adjusted covariates and exposure time window (Figure 2). For example, the unadjusted model reported the estimated effect as a birthweight reduction of 0.84 g (95% CI: −0.53 to 2.22). The lag-specific associations did not display a clear time-varying pattern (Figure S1), which suggests a linear increase in strength in the association with increased length of the exposure time window (Figures S2 & 2). The subpopulation-specific results suggest that the association was potentially heterogeneous (Figure S3). The association was stronger for female infants, newborns of nulliparous mothers, or newborns of unemployed mothers, compared with the corresponding references (i.e., male infants, newborns of multiparous mothers, or newborns of employed mothers, respectively). The nonlinear analysis confirmed the adverse effect of LFS on birthweight (Figure 3). The sublinear association suggests a saturated effect of a ∼60 g birthweight reduction for exposure to extreme fires, which contributed to a PM_2.5_ concentration > ∼20 µg/m^3^.

**Figure 2.**
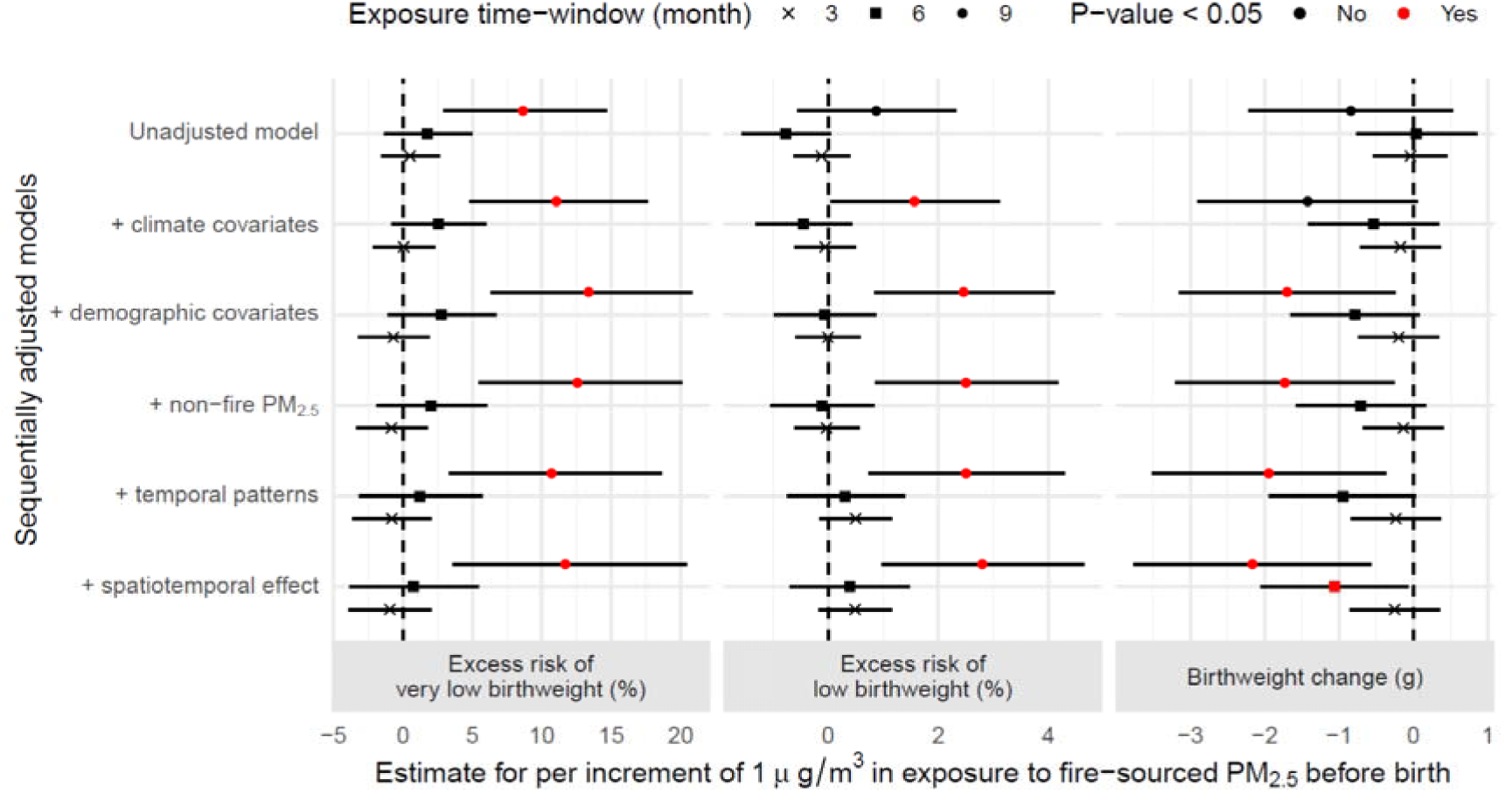
Estimated associations between gestational exposure to fire-sourced PM_2.5_ and birthweight change, low birthweight, or very low birthweight. The dots represent the point-estimates, and bars represent the corresponding 95% confidence intervals.

**Figure 3.**
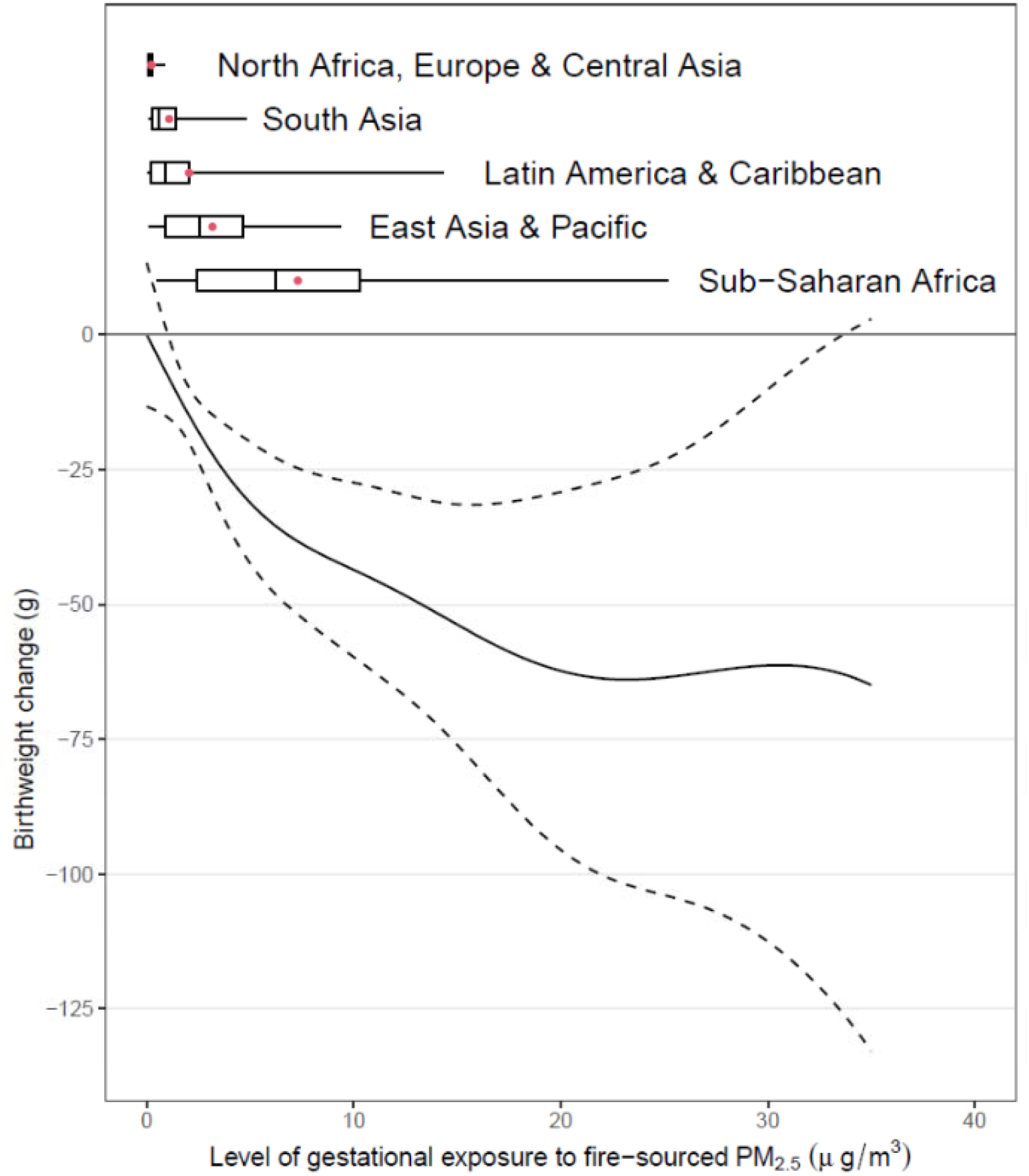
The nonlinear association between gestational exposure to fire-sourced PM_2.5_ and change in birthweight. The solid line represents the point-estimates, the dashed line represents the 95% confidence intervals, the boxplots represent the distributions of different exposure levels by regions; and the red dots represent the mean exposure level.

### Association between LFS and VLBW

Our fully adjusted models suggested that LFS exposure during the 9 months before birth was significantly associated with an increased risk of LBW or VLBW. According to the estimates, the risks of LBW and VLBW increased by 2.80% (95% CI: 0.97–4.66) and 11.68% (95% CI: 3.59–20.40), respectively, for each 1 µg/m^3^ increase in exposure to fire-sourced PM_2.5_. Compared to LBW, VLBW was more strongly associated with fire-sourced PM_2.5_. The estimated association for LBW was sensitive to different covariate adjustments (Figure 2) and was potentially heterogeneous among subpopulations (Figure S3). The association for VLBW was robust, given the adjustments for different covariates (Figure 2), and was less heterogeneous (Figure S3). Among all effect modifiers, only the type of cooking energy, which was indicated for indoor sources of particulate matter, significantly changed the association between VLBW and fire-sourced PM_2.5_ (P-value = 0.02, Figure S3). The use of unclear cooking energy, as a competing risk factor for fire-sourced PM_2.5_, significantly weakened the association between VLBW and fire-sourced PM_2.5_.

To further explore why VLBW was more strongly associated with fire-sourced PM_2.5_ than was LBW, we developed a model to estimate the baseline-varying association between birthweight and fire-sourced PM_2.5_. As a result, the absolute change in birthweight for per unit exposure to fire-sourced PM_2.5_ varied with the baseline of family-level mean birthweight (Figure S4). Samples with an extremely low or high baseline birthweight were more susceptible to fire-sourced PM_2.5_ than were those with a moderate baseline birthweight, in terms of the absolute birthweight change (Figure S4). As a reduction in birthweight and a family-level baseline co-determined the increased incidence of LBW or VLBW, the relative change in birthweight reflected the relevant risk. Newborns from families with a lower baseline were more susceptible to fire-sourced PM_2.5_, in terms of a relative birthweight reduction (Figure 4). This finding partially explains why gestational exposure to fire-sourced PM_2.5_ was robustly and strongly associated with VLBW. According to our estimates, a 1 µg/m^3^ increase in fire-sourced PM_2.5_ was associated with a relative birthweight reduction of 1.47% (95% CI: 0.06– 2.88), 0.54% (95% CI: 0.03–1.05), 0.16% (95% CI: 0.01–0.31), or 0.03% (95% CI: −0.04 to 0.10) for samples with a baseline birthweight of 1,500, 2,000, 2,500, or 3,000 g, respectively.

**Figure 4.**
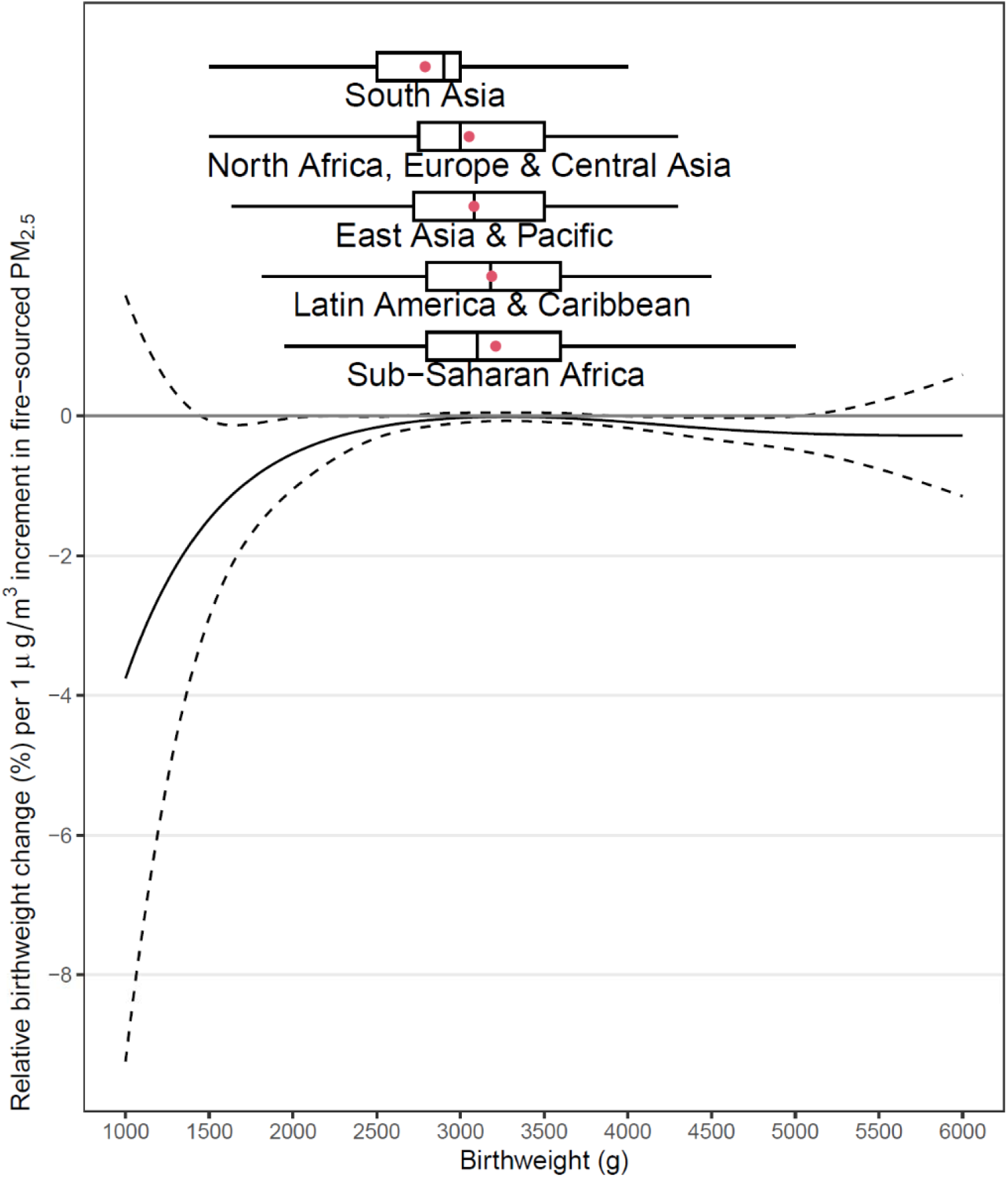
Baseline-varying association between gestational exposure to fire-sourced PM_2.5_ and relative change in birthweight. The solid line represents the point-estimates, the dashed line represents the 95% confidence intervals, and the boxplots represent the distributions of different birthweights by region.

### Sensitivity analyses

First, wet explored potential recall bias using sensitivity analyses, and the estimated associations were not significantly different according to recall period (Figure S3). Second, we examined the spatial variations in the estimated associations. Although analysis of differences in region-specific associations reported non-significant results (Figure S3), we could not completely rule out the possibility of spatial heterogeneity in the estimated results. As most of our analyzed samples were from Sub-Saharan Africa, only those estimates were significant, and the remaining region-specific results had wide-ranging 95% CIs (Figure S3). Third, we explored whether the estimated adverse effects were attributable to ambient LFS exposure or other impacts (e.g., social stress) relevant to landscape fires. The estimated effects of exposure to fire-sourced PM_2.5_ were not significantly different between locations without burning and those with burning (Figure S3). These results confirm that ambient exposure was the major exposure pathway for our analyzed outcomes.

## Discussion

This study revealed an association between increased risk of VLBW and exposure to LFS during pregnancy in 54 LIMCs. We found that gestational exposure to fire-sourced PM_2.5_ potentially reduced birthweight, and the infants with a low family-level mean of birthweight were susceptible to the effect of LFS. We present the first sibling-matched study on the association between birthweight and fire-sourced PM_2.5_ from multiple LIMCs.

LFS contains numerous hazardous pollutants and exerts both short- and long-term effects on human health^34^. An adverse birth outcome is one of the consequences. Holstius et al.^23^ estimated that the mean birthweight of fire-exposed infants are 7.0 g (95% CI: 11.8–2.2), 9.7 g (95% CI: 14.5–4.8), and 3.3 g (95% CI: −0.6 to 7.2) lower than those of unexposed infants when wildfires occurred during the first, second, and third trimesters, respectively. In a retrospective cohort study in Brazil, fire-exposed mothers (defined as the highest PM_2.5_ concentration quartile) reported a higher risk of LBW than did unexposed mothers. The adjusted odds ratios for exposure during the second and third trimesters were 1.51 (95% CI: 1.04–2.17) and 1.50 (95% CI: 1.06–2.15), respectively^24^. Abdo et al.^25^ showed that a 1 μg/m^3^ increase in the wildfire-sourced PM_2.5_ concentration during the first trimester was associated with a 5.7 g (95% CI: 0.4–11.1) reduction in birthweight, in United States. In contrast, an Australian cross-sectional study reported that the average birthweight of male infants exposed to the 2003 Canberra wildfires was significantly higher than that of unexposed peers^26^. That study attributed the positive association between wildfire smoke and birthweight change to the potentially increased levels of maternal blood glucose after exposure.^26^ The mixed results might be due to differences among studies in terms of epidemiological design, population characterization, exposure time window, and competing risk factors. Our results are comparable with most previous findings. Additionally, although there are few studies on fire-sourced PM_2.5_ similar to ours, a large number of studies have focused on the effect of gestational exposure to urban ambient PM_2.5_ on birthweight. A recent meta-analysis, which included more than 2.7 million participants, reported that the pooled risk ratio of LBW for a 10 μg/m^3^ increase in PM_2.5_ exposure was 1.081 (95% CI 1.043–1.120)^21^. Considering the abundance of toxic organic particles derived from biomass burning, the association of birthweight with fire-sourced PM_2.5_ should be stronger than that with urban PM_2.5_.

The family-level mean birthweight varied widely between households (Table 2). These variations may have been caused by many factors, including genetics, socioeconomic position (e.g., nutrition), and indoor household pollution. As birthweight is positively correlated with gestation length, a poor family-level baseline birthweight comprehensively indicates a combined risk of short gestation and LBW. Because the cumulative effect of fire-sourced PM_2.5_ was observed to increase with exposure duration (Figure S2), a larger birthweight at the family level, as indicated for a longer gestation, increased the absolute effect of the per-unit LFS exposure (Figure S4).

As our models did not control for gestation length due to the lack of relevant data, the estimated associations reflected the joint outcome of LFS exposure by reducing gestation length or birthweight. The fetus grows in a sublinear temporal pattern^35^ during the late gestational stage, which suggests its weight increases to an approximately stable level before birth. Considering this sublinear relationship and given the different baseline birthweights, the same reduction in gestation length led to different reductions in birthweight. The marginal effect of a shorter gestation period on the reduction in birthweight is larger for an infant with a lower baseline birthweight. For instance, if an infant is expected to reach the stable level of fetal weight, a marginal reduction in its gestational age will not considerably change the birthweight. As exposure to LFS has been reported to cause a short gestation^25^, its indirect effect on reduced birthweight can be enhanced by poor baseline maternal health.

Reducing the risk of LBW is one of the 2025 WHO global targets, and it contributes to achieving the Sustainable Development Goals. Although major interventions to reduce LBW have focused on improving maternal nutrition, environmental toxic pollutants, also identified as risk factors, can additionally contribute to the relevant disease burden. For instance, exposure to air pollution was linked to a global burden of 476,000 infant deaths in 2019 during the first month of life via increased risks of LBW and premature birth^36^. LFS induces peak concentrations of PM_2.5_ lasting for a few weeks or months. Unlike other environmental exposures, such as urban PM_2.5_, LFS exposure is unequally distributed spatially and temporally. The individual-level impacts on exposed mothers are not negligible. Particularly, LFS exposure frequently occurred in LMICs (Figure 1), where the fire-sourced PM_2.5_ adversely affected infants with other risk factors, such as malnutrition. However, due to the lower specificity of our model evaluating birthweight without adjusting for gestation length, we could not evaluate the clinical outcomes or disease burden of the LFS-attributed birthweight reductions. As there is an increasing probability of landscape fire under global climate change, assessing the adverse impacts of LFS exposure is of public health importance to protect maternal health in LMICs. Future studies should be conducted to answer this question.

This study was associated with the following limitations. First, the lack of data on specific gestation lengths in the DHSs limited our exploration. Without this information, we could not distinguish the exposure time window, which introduced potential exposure misclassifications into the epidemiological analyses. Also, we could not determine the clinical importance of the analyzed birthweight without normalizing it to gestation length. Second, characterizing LFS exposure is always challenging. Previous studies measured exposure using surrogates, such as the temporal duration of landscape fires or satellite remote sensing of fire points. To increase interpretability, we utilized fire-sourced PM_2.5_ as a direct indicator of ambient exposure to LFS. However, as the exposure assessments were based on a CTM, the uncertainties introduced by the modeling procedure could not be avoided completely. The potential errors enhanced the exposure misclassifications in this study and thus may have resulted in a biased association. Third, since the analyzed samples were living newborns, our estimates were potentially subjected to survival bias. For instance, fire-sourced PM_2.5_ was associated with extremely reduced birthweight, which increases the probability of stillbirth or neonatal death. In other words, our sample may have included fewer embryos more susceptible to fire-sourced PM_2.5_ than expected. Therefore, survival bias could have led to an underestimated association by ignoring those susceptible embryos. Finally, a sibling-matched study requires a selected sample, which may have lowered the representativeness of this study. The inclusion of infants of the DHS in this study was determined by many factors, such as family size and planning, willingness to bear a child, socioeconomic position, and contraceptive behavior. However, some of those factors could not be easily measured, which impeded the development of a set of sampling weights to increase the representativeness of our estimates.

## Conclusions

Using a global-scale sibling-matched case–control study, an association was identified between gestational exposure to fire-sourced PM_2.5_ and reduced birthweight in LMICs. The association was sensitive to regression model settings and potential heterogeneity among different subpopulations. Newborns from families with a low baseline level of birthweight were susceptible to LFS-associated birthweight reduction, which was consistent with our finding of a strong and robust association between fire-sourced PM_2.5_ and VLBW. This study adds to the epidemiological evidence on the adverse effects of LFS on birthweight. Relevant interventions regarding frequently occurring landscape fires, such as climate change mitigations, should be taken to protect maternal and infant health.

## Data Availability

All analyses in this paper are based on open-accessed data.

