## Supplemental materials for "Exposure to landscape fire smoke extremely reduced birthweight in low- and middle-income countries"

***Supplemental text***

GEOS-Chem is a widely-utilized and well-designed global CTM to simulate the chemical components of atmosphere. This study used the 11-01 version of GEOS-Chem model. The meteorological fields to drive the CTM were directly obtained from an assimilated dataset, Modern-Era Retrospective analysis for Research and Applications Version 2 (MERRA-2), which is maintained by the Global Modeling and Assimilation Office of National Aeronautics and Space Administration (NASA), United States (US). MERRA-2 has a spatial resolution of 0.5° × 0.625°, and is freely distributed by the Goddard Earth Sciences Data and Information Services Center of US NASA (https://disc.gsfc.nasa.gov/).

For the chemical reactions, our GEOS-Chem model applied the full O_x_−NO_x_−CO−VOC−HO_x_ mechanism. The model simulated the reactions relevant to multiple atmospheric components, such as sulfate-nitrate-ammonium, primary and secondary carbonaceous aerosols, mineral dusts and sea-salts. For instance, we simulated the interactions of sulfate-nitrate-ammonium, according to the ISOROPIA-II thermo-dynamical equilibrium^37^. Since this study was focused on PM_2.5_ simulations, we evaluated the aerosol results extensively using measurement data ^38-40^. In the model, anthropogenic emissions, from 2000 to 2014, were obtained from the global inventory of Community Emissions Data System (CEDS) ^41^, the fire emissions during the same period, were collected from the Global Fire Emission Database (GFED4s). We ran the GEOS-Chem simulations with a six-month spin-up starting from July 1999, with a horizonal resolution of 2° × 2.5°, and a vertical resolution of 47 layers. We took the PM_2.5_ in the bottom layer as the ground-surface concentrations of PM_2.5_.

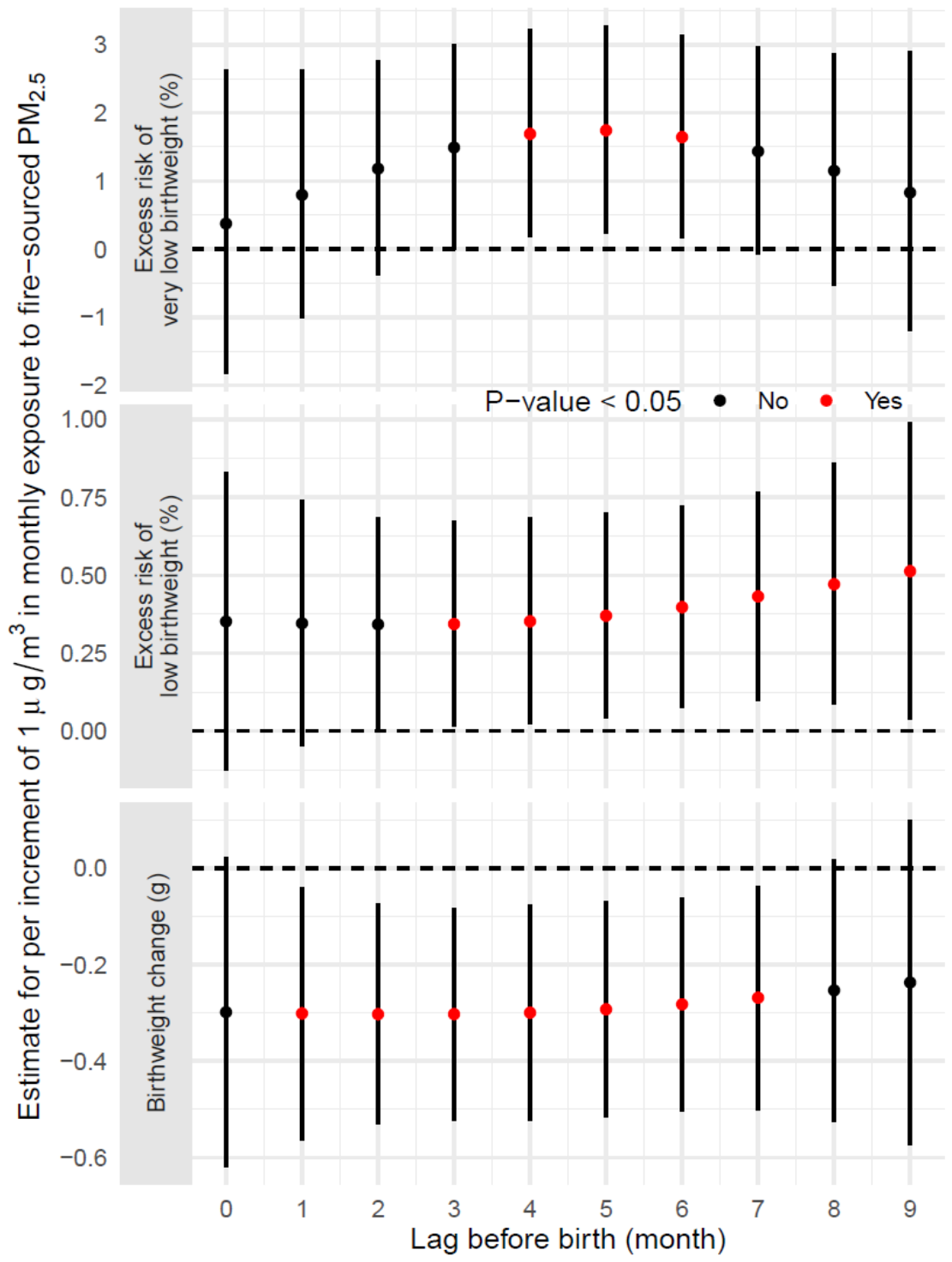

Figure S 1 The estimated associations between fire-sourced PM_2.5_ and birthweight change, low birthweight or very low birthweight, by different lags. The n^th^ lagged exposure is defined as the concentration of fire-sourced PM_2.5_ during the n^th^ month before birth. The results are estimated from the lag-distributed models.

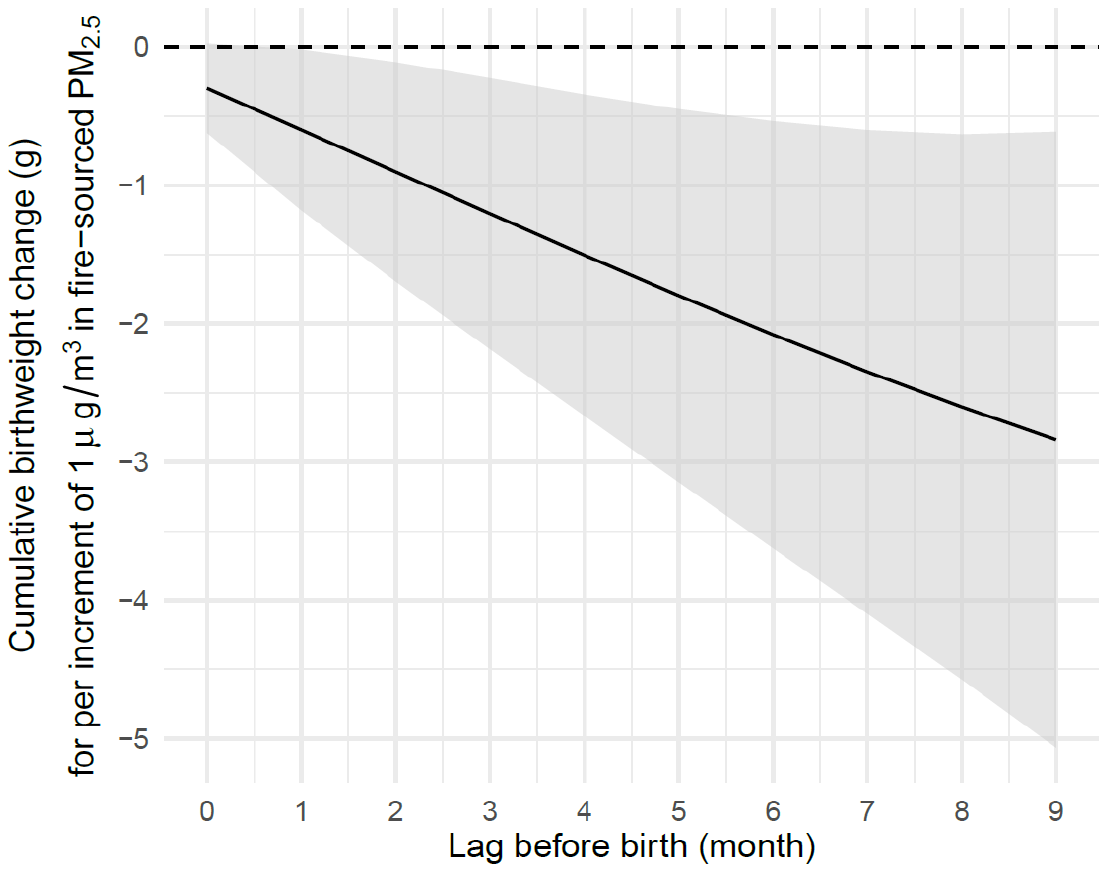

Figure S 2 The cumulated birthweight change associated with an average of fire-sourced PM_2.5_ concentrations from a lagged month to birth.

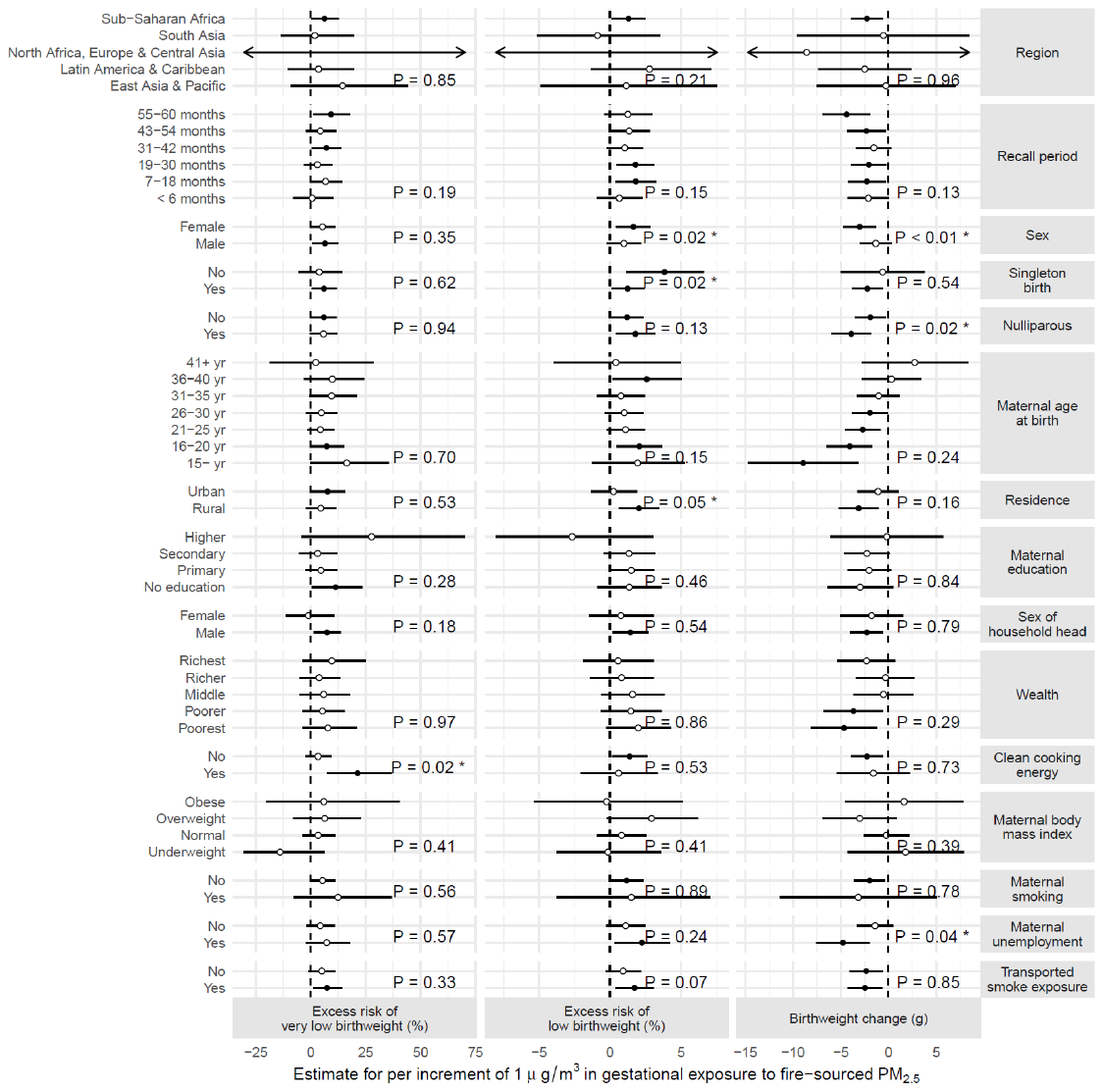

Figure S 3 The subpopulation-specific associations between gestational exposure to fire-sourced PM_2.5_ and birthweight change, low birthweight and very low birthweight.

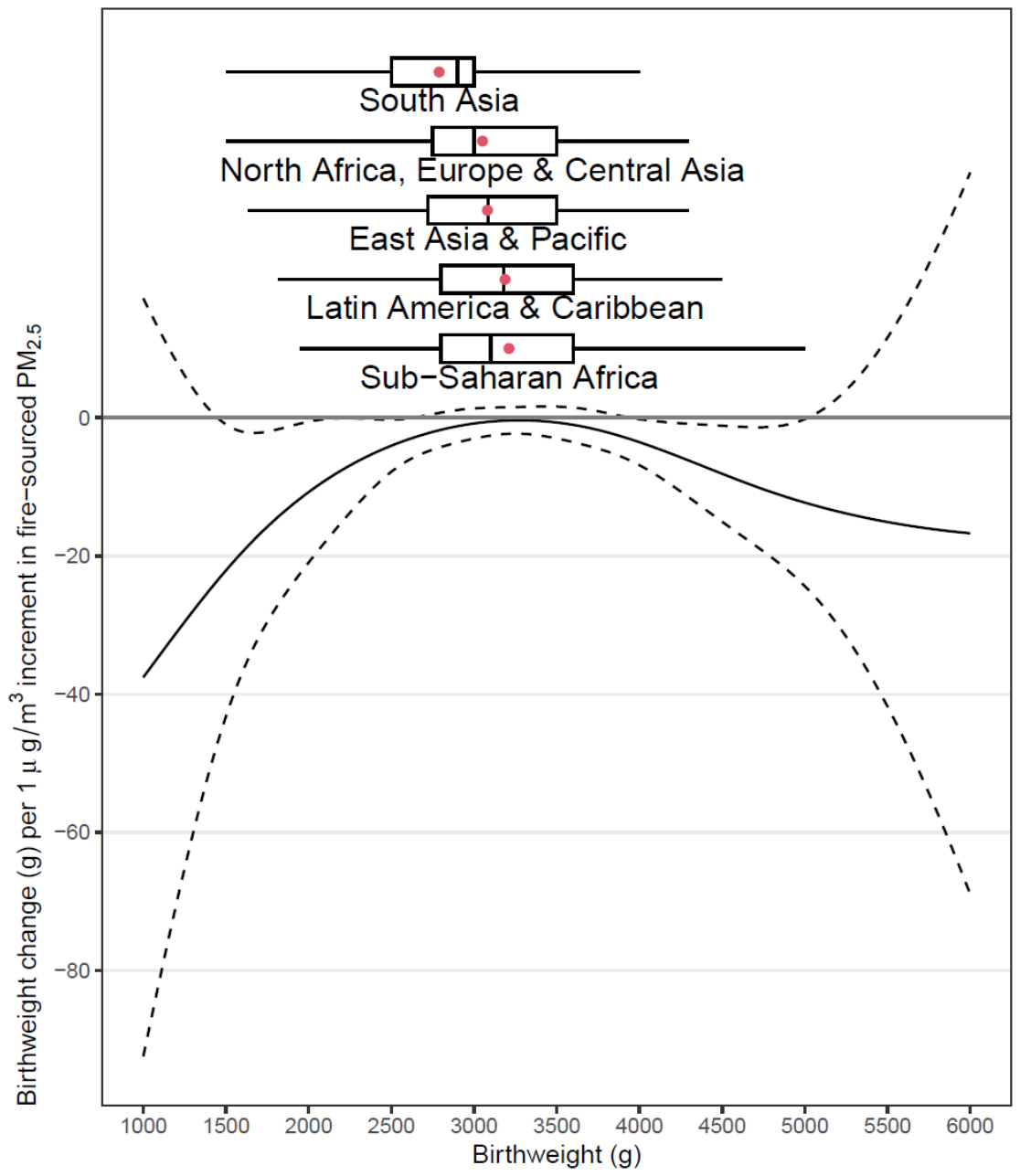

Figure S 4 The baseline-varying association between gestational exposure to fire-sourced PM_2.5_ and absolute birthweight change. The solid line presents the point-estimates, the dashed line presents the 95% confidence intervals, and the boxplots present the distributions of different birthweight levels by regions.

Table S1 Population characteristics.

| Variable | Group | All samples | low-birthweight births | very-low-birthweight births |
| --- | --- | --- | --- | --- |
| Categorical variable | | N (percentage, %) | | |
| Total | | 227,948 (100) | 31,854 (100) | 2,912 (100) |
| Sex | Male | 114,772 (50.4) | 14,840 (46.6) | 1,379 (47.4) |
|  | Female | 113,176 (49.6) | 17,014 (53.4) | 1,533 (52.6) |
| Year of birth | 2000 | 1,169 (0.5) | 166 (0.5) | 15 (0.5) |
|  | 2001 | 4,366 (1.9) | 645 (2.0) | 64 (2.2) |
|  | 2002 | 5,837 (2.6) | 777 (2.4) | 90 (3.1) |
|  | 2003 | 7,853 (3.4) | 959 (3.0) | 97 (3.3) |
|  | 2004 | 9,729 (4.3) | 1,055 (3.3) | 114 (3.9) |
|  | 2005 | 10,091 (4.4) | 1,136 (3.6) | 113 (3.9) |
|  | 2006 | 13,222 (5.8) | 1,514 (4.8) | 120 (4.1) |
|  | 2007 | 13,806 (6.1) | 1,697 (5.3) | 164 (5.6) |
|  | 2008 | 15,324 (6.7) | 1,903 (6.0) | 153 (5.3) |
|  | 2009 | 18,004 (7.9) | 2,034 (6.4) | 183 (6.3) |
|  | 2010 | 25,159 (11.0) | 3,305 (10.4) | 295 (10.1) |
|  | 2011 | 29,397 (12.9) | 4,537 (14.2) | 391 (13.4) |
|  | 2012 | 27,284 (12.0) | 4,352 (13.7) | 397 (13.6) |
|  | 2013 | 23,015 (10.1) | 3,920 (12.3) | 388 (13.3) |
|  | 2014 | 23,692 (10.4) | 3,854 (12.1) | 328 (11.3) |
| Singleton (n = 1) or multiple births (n > 1) | n = 1 | 211,276 (92.7) | 23,910 (75.1) | 1,740 (59.8) |
|  | n = 2 | 8,268 (3.6) | 3,876 (12.2) | 552 (19.0) |
|  | n = 3 | 8,276 (3.6) | 3,962 (12.4) | 573 (19.7) |
|  | n = 4 | 124 (0.1) | 102 (0.3) | 45 (1.5) |
|  | n = 5 | 4 (0.0) | 4 (0.0) | 2 (0.1) |
| Region | East Asia & Pacific | 10,059 (4.4) | 1,259 (4.0) | 173 (5.9) |
|  | Latin America & Caribbean | 25,747 (11.3) | 3,040 (9.5) | 372 (12.8) |
|  | North Africa, Europe & Central Asia | 21,932 (9.6) | 2,948 (9.3) | 404 (13.9) |
|  | South Asia | 57,004 (25.0) | 11,307 (35.5) | 981 (33.7) |
|  | Sub-Saharan Africa | 113,206 (49.7) | 13,300 (41.8) | 982 (33.7) |
| Recall period (years, from birthdate to survey date) | < 0.5 | 22,628 (9.9) | 2,586 (8.1) | 218 (7.5) |
|  | 0.5~1.5 | 40,984 (18.0) | 5,421 (17.0) | 496 (17.0) |
|  | 1.5~2.5 | 48,707 (21.4) | 7,258 (22.8) | 674 (23.1) |
|  | 2.5~3.5 | 40,755 (17.9) | 6,086 (19.1) | 600 (20.6) |
|  | 3.5~4.5 | 52,658 (23.1) | 7,442 (23.4) | 647 (22.2) |
|  | 4.5~5 | 22,216 (9.7) | 3,061 (9.6) | 277 (9.5) |
| Birth order | 1 | 47,969 (21.0) | 7,737 (24.3) | 767 (26.3) |
|  | 2 | 70,468 (30.9) | 9,765 (30.7) | 864 (29.7) |
|  | 3 | 41,015 (18.0) | 5,467 (17.2) | 477 (16.4) |
|  | 4 | 25,521 (11.2) | 3,324 (10.4) | 302 (10.4) |
|  | 5 | 16,517 (7.2) | 2,103 (6.6) | 190 (6.5) |
|  | 6 | 10,813 (4.7) | 1,406 (4.4) | 132 (4.5) |
|  | 7 | 15,645 (6.9) | 2,052 (6.4) | 180 (6.2) |
| Maternal age | < 16 | 1,712 (0.8) | 315 (1.0) | 41 (1.4) |
|  | 16~20 | 43,166 (18.9) | 6,895 (21.6) | 650 (22.3) |
|  | 21~25 | 82,870 (36.4) | 11,666 (36.6) | 1,056 (36.3) |
|  | 26~30 | 56,738 (24.9) | 7,317 (23.0) | 628 (21.6) |
|  | 31~35 | 28,694 (12.6) | 3,591 (11.3) | 345 (11.8) |
|  | 36~40 | 12,210 (5.4) | 1,706 (5.4) | 163 (5.6) |
|  | > 40 | 2,558 (1.1) | 364 (1.1) | 29 (1.0) |
| Residence | Rural | 142,202 (62.4) | 20,396 (64.0) | 1,740 (59.8) |
|  | Urban | 85,746 (37.6) | 11,458 (36.0) | 1,172 (40.2) |
| Education | No education | 52,061 (22.8) | 8,436 (26.5) | 691 (23.7) |
|  | Primary | 74,510 (32.7) | 9,476 (29.7) | 855 (29.4) |
|  | Secondary | 82,914 (36.4) | 11,600 (36.4) | 1,110 (38.1) |
|  | Higher | 18,455 (8.1) | 2,340 (7.3) | 256 (8.8) |
|  | Unknown | 8 (0.0) | 2 (0.0) | 0 |
| Head of household | Female | 37,259 (16.3) | 5,309 (16.7) | 499 (17.1) |
|  | Male | 190,689 (83.7) | 26,545 (83.3) | 2,413 (82.9) |
| Wealth level | Richest | 41,464 (18.2) | 4,948 (15.5) | 471 (16.2) |
|  | Richer | 45,145 (19.8) | 6,091 (19.1) | 567 (19.5) |
|  | Middle | 47,235 (20.7) | 6,567 (20.6) | 591 (20.3) |
|  | Poorer | 47,539 (20.9) | 6,920 (21.7) | 609 (20.9) |
|  | Poorest | 46,231 (20.3) | 7,224 (22.7) | 661 (22.7) |
|  | Unknown | 334 (0.1) | 104 (0.3) | 13 (0.4) |
| Type of cooking energy | Unclean | 160,760 (70.5) | 21,897 (68.7) | 1,780 (61.1) |
|  | Clean | 53,348 (23.4) | 7,713 (24.2) | 869 (29.8) |
|  | Unknown | 13,840 (6.1) | 2,244 (7.0) | 263 (9.0) |
| Body mass index level | Underweight | 20,756 (9.1) | 4,154 (13.0) | 327 (11.2) |
|  | Normal | 97,169 (42.6) | 13,863 (43.5) | 1,164 (40.0) |
|  | Overweight | 32,759 (14.4) | 4,006 (12.6) | 397 (13.6) |
|  | Obese | 14,265 (6.3) | 1,776 (5.6) | 224 (7.7) |
|  | Unknown | 62,999 (27.6) | 8,055 (25.3) | 800 (27.5) |
| Smoking | No | 206,937 (90.8) | 28,629 (89.9) | 2,599 (89.3) |
|  | Yes | 9,105 (4.0) | 1,470 (4.6) | 132 (4.5) |
|  | Unknown | 11,906 (5.2) | 1,755 (5.5) | 181 (6.2) |
| Unemployed mother | No | 102,861 (45.1) | 11,874 (37.3) | 1,056 (36.3) |
|  | Yes | 76,781 (33.7) | 10,501 (33.0) | 1,067 (36.6) |
|  | Unknown | 48,306 (21.2) | 9,479 (29.8) | 789 (27.1) |
| Continuous variable | | Mean (standard deviation) | | |
| Birthweight (g) | | 3081.91 (724.11) | 1962.22 (368.99) | 1092.79 (222.60) |
| Fire-sourced PM_2.5_ (µg/m^3^) | | 4.29 (5.53) | 3.35 (4.65) | 2.64 (3.86) |
| Non-fire-sourced PM_2.5_ (µg/m^3^) | | 37.43 (29.60) | 42.03 (31.90) | 40.99 (32.61) |
| Temperature (K) | | 296.38 (4.78) | 296.90 (4.37) | 296.78 (4.26) |
| Humidity (g/kg) | | 12.36 (3.51) | 12.18 (3.40) | 12.00 (3.56) |
| Maternal age (year) | | 25.52 (5.71) | 25.13 (5.79) | 25.05 (5.89) |
| Number of matched siblings | | 2.13 (0.36) | 2.18 (0.43) | 2.26 (0.53) |
